# Manifestations of mortality based global data of COVID-19; unifying global model through single parameter

**DOI:** 10.1101/2020.05.13.20100255

**Authors:** V.K. Jindal

## Abstract

Critical inspection of the world data of COVID-19 mortality rates per population number has been made and used to express extensive variations in mortality over the globe in terms of a powered parameter λ varying from 0 to 1.2 expressed as a measure of strength of primary infection, originating from China source. The copying process is degenerating successively while infection is passed on to secondary subjects. We have been able to correlate global data through this parameter; any value close to or less than 1 shows significant impact of diluted multiple secondary effect. Further, the scatter diagram shows no effect of temperature of the geographical location and so is likely as the virus is only being spread from either contact or close proximity – the virus does not need to face highs and lows of temperatures of the environment. It stays only in the range of human body temperature and appears to be stable in 36–40°C range. If it faces the environmental temperatures it is possible for its quicker deactivation but that situation never arises for this virus except when it spreads from surfaces.

---

COVID-19 disease has been in focus for over 5 months now and has transformed the world into a complete rethink on technical, economic, political, philosophical and medical fronts. The numbers of persons infected and mortality are alarming to say the least. They are a challenge to our scientific understanding and indeed also leading us to rediscover our philosophical thinking raising questions unconnected to science drawing inferences on whether the uniformity and equality we as scientists and technocrats are trying to deliver to our masses all over the globe serve the purpose we thought it would. The discriminatory nature of the mortality data from geographical position point of view also puzzles us and we must be exploring the data more realistically because data is one important truth before us. We therefore use the data to postulate what is possible at this point of time for the sake of exploiting new thinking strategies.

Fortunately, enormous world data related to COVID-19 has been uploaded publicly by John Hopkins University, coronavirus resource centre [1]. This data is highly revealing, the mortality varying from some 75 per 100,000 of population (Belgium) to nearly 0.15 per 100,000 of population (India), or even lower for some small Countries. This variation has a message and we must read it.

We read it combining it with some facts. Firstly, we notice that the disease originated in Wuhan in China last year end. It slowly and stealthily overtook the whole globe. Why this variation in quick time of a few months is the question that bothers us. It is apparent that there is a great influence of Chinese tourists and vice versa (they visiting them) who frequented most of the Countries having a higher mortality. The influence on many such populations who got secondary infections do not have such high mortality. That indicates to the fact that the virus multiplication is attenuated while the infection is carried forward from originally infected to subsequently infected, like an analogue of non-digital photocopy, digital copy being reserved for primary infection sources. We know such analogue copying machine reduces the image by multiplying fraction successively.

Secondly, a fact we notice that the influence of geographical local temperature when plotted against the mortality rate only returns a random scatter diagram hardly showing any correlation to high and low temperature bias. This is truly a person to person dominant infection and the only temperature that matters to the virus is human body temperature indicating thereby that the virus is very active in the range around 37°C, even surviving under fever. Therefore, there is no deactivation possibly in the range 36–40°C. If at all there is some role of higher temperature deactivation, it will manifest when infection spreading through surfaces is accounted for and that component needs to be worked out.

Although a model for India specific disease was proposed earlier [2], we would like to generalize it by introducing an origin (China) factor λ to couple with the usual growth rate *R*_0_, such that the growth eq for the number of infected persons is now governed by χ instead (compare with Eq. 8–10 of [2])

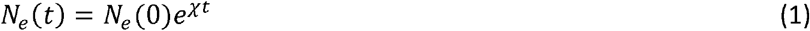

And for *t* > *t*_0_, (*t*_0_ is the onset of lockdown time)

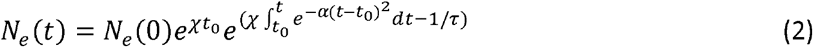

Which is solved as,

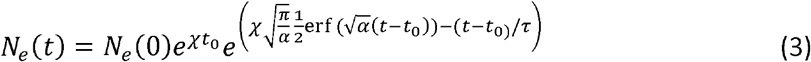

Where χ = λ*R*_0_. Time *t*_0_ is the time (day of onset of lockdown announcement), α dictates the width of the decay of χ and τ dictates the mean recovery time. The *R*_0_ used by us is the effective growth rate of an exponential least square fit to pre-lockdown period

Dividing the global population into different zones dictated by choosing λ as 1, 1.1 and 1.2 with λ = 1 taken for India specific data, we re-calculate the growths of various numbers. The effect of primary infection (λ = 1.2) makes enormous difference in mortality numbers as shown in Fig. 1. It is desirable to choose country specific lockdown onset time and recovery pattern to arrive at full implications as applicable to all numbers. The Countries Germany, Iran, Canada, Portugal, Ecuador, Austria and Denmark can be categorised under category λ = 1.1. Similarly, Italy, USA, United Kingdom, Spain, France, Belgium and 8 other Countries fall in λ = 1.2 category. Indeed, these three values are just to divide in low, middle and high range. A country specific value can be chosen.

**Figure 1.**
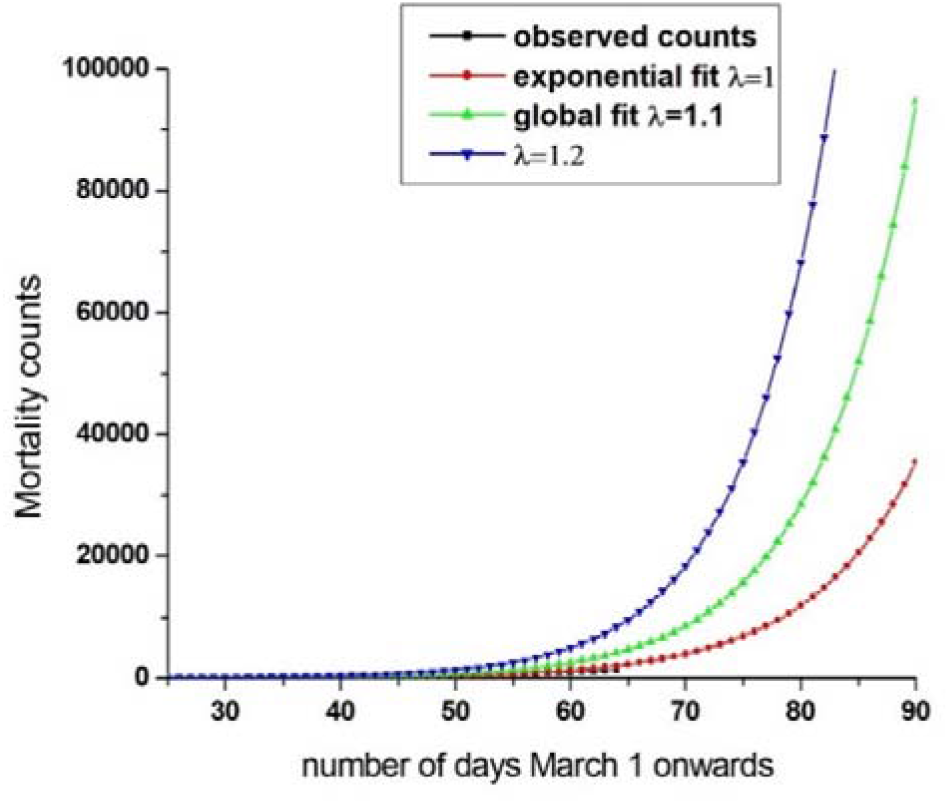
Mortality data (black dots) of India specific disease and its fit. This assumes λ = 1. The global data is speculated to be derived from λ varied or raised by upto 20% in λ value. The observed death numbers for India is black points curve hidden under the fit in red.

With new expressions provided, coupled with Gaussian *R*_0_, forecasting models for any Country can be formulated.

There are other interesting models which also suggest different ideas [3–6] on modelling the available COVID-19 data. The inference drawn in [6] from data about role of temperature has been mentioned contrary to our findings. Collecting mean temperature during March-April period of various COVID affected global regions, where mortality data has been provided by JH University [1], we have tried to establish any correlation if the mortality per population is following some pattern with mean temperature. The scatter plots for all data, and parts of the data has been presented in Figs. 2–5.

**Figs 2(left) and 3 (right).**
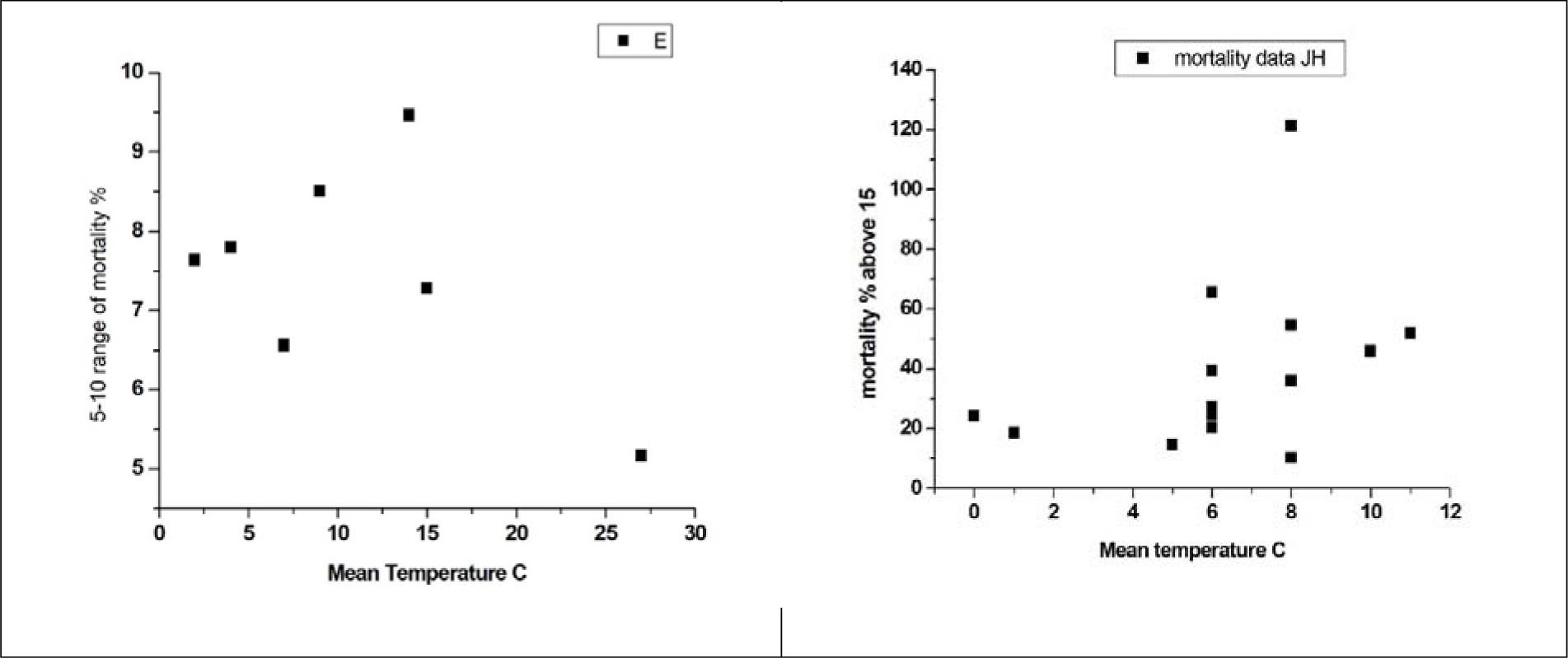
Showing Countries with mortality rates between 5-10% per 100K of population and above 15% of mortality, respectively

**Figure 4.**
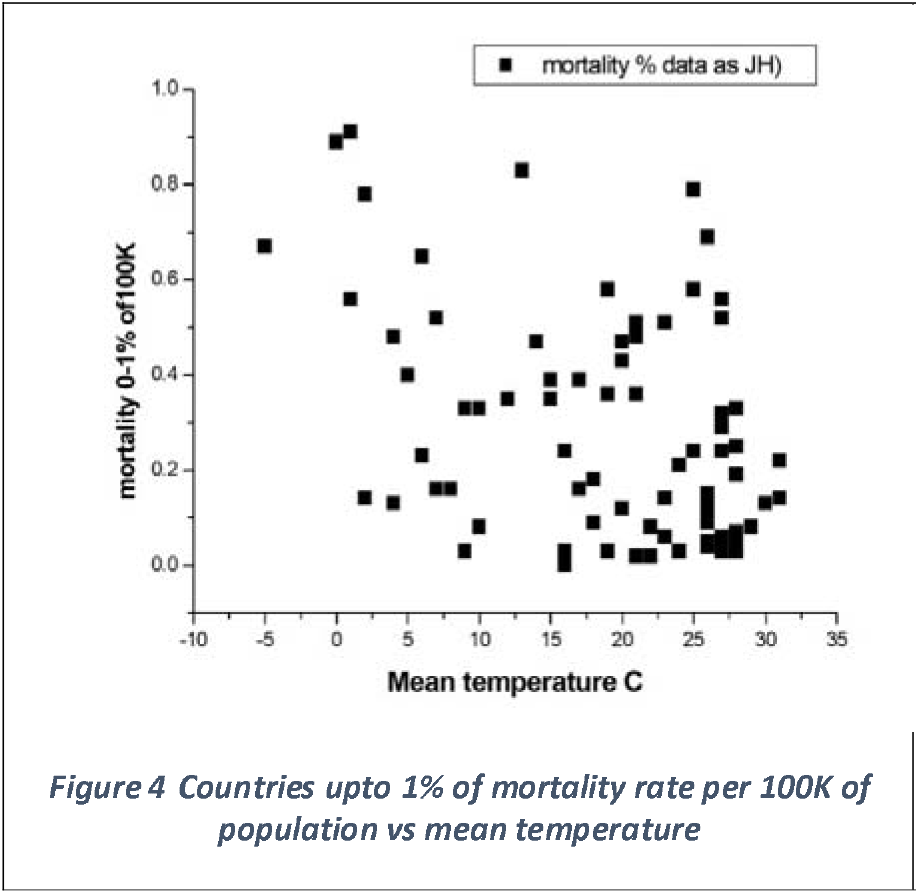
Countries upto 1% of mortality rate per 100K of population vs mean temperature

Whereas Fig 5 shows all countries together and leads to a misleading conclusion that majority of the data can be linearly fitted to a straight line which might show vanishing mortality around 55°C. This becomes evident from the data shown separately of different mortality rates as in Fig 2 to 4. All data show no real pattern with temperature and no worthwhile conclusion can be drawn.

**Figure 5.**
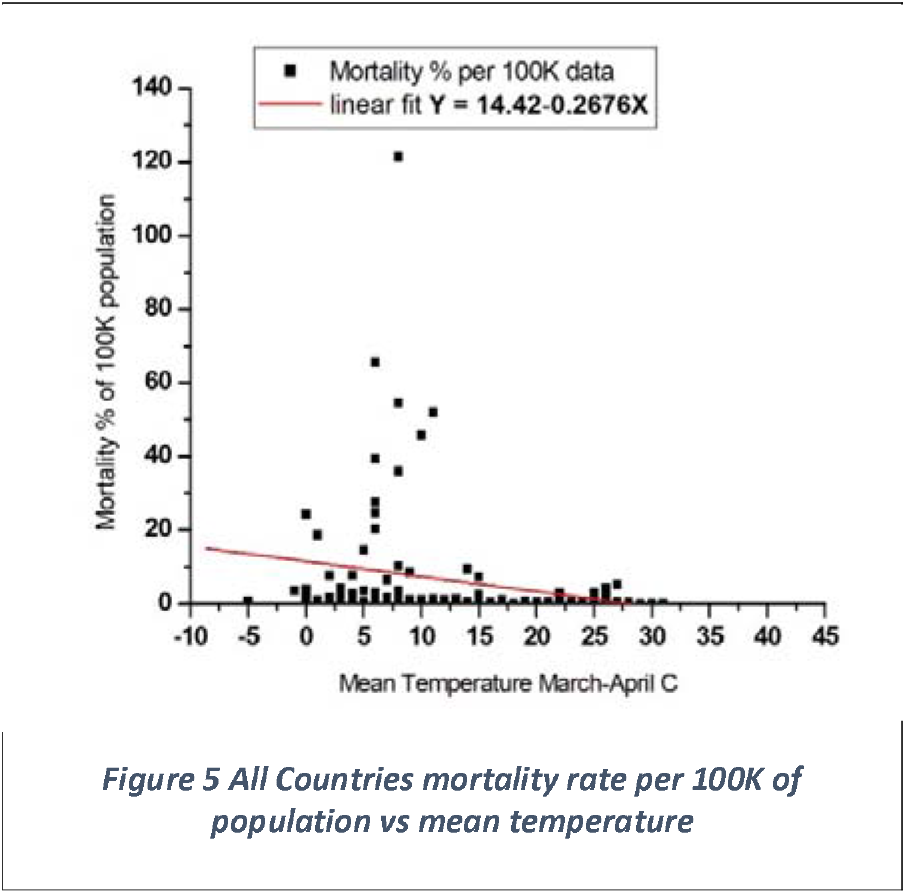
All Countries mortality rate per 100K of population vs mean temperature

In conclusion, we would like to reemphasize that the COVID-19 disease can be classified as primary infection with λ raised to 1.2 from India specific value of 1. Various International cities have different impact expressible in terms of λ, the China factor where the population has direct impact of tourism from infected parts of China. When the infection happens to be caused by secondary infection, there is considerable subdued impact on the severity.

Further, temperature of the environment has no impact as long as the virus is transferred from contact or close proximity because it very well survives human body temperature. The effect of high climatic temperature would be expected if the infection becomes dominant from surfaces or environment resident virus.

## Data Availability

Global data resource centre https://coronavirus.jhu.edu/data

## Acknowledgements

The author highly appreciates the help provided by my son, Dr. Prashant Jindal (University Institute of Engineering & Technology, Panjab University, Chandigarh) for collecting large part of the information and data and bringing out the temperature scatter, leading me to conclude non correlation to the climatic temperature.

